# Preferences for the management of sexually transmitted infections in the South African health system: A discrete choice experiment

**DOI:** 10.1101/2022.03.07.22271994

**Authors:** Collins C Iwuji, Catherine E Martin, Diantha Pillay, Patience Shamu, Susan Nzenze, Mercy Murire, Laura Ashleigh Cox, Alec Miners, Carrie Llewellyn, Saiqa Mullick

## Abstract

**Introduction:** Young people have a disproportionate burden of sexually transmitted infections. Despite strengthening HIV prevention with the introduction of PrEP, STI services have remained relatively unchanged, and the standard of care remains syndromic management. We used a discrete choice experiment to investigate young people’s preferences for the diagnosis and treatment of STIs in South Africa.

**Methods and Findings:** Between 1 March 2021 and 20 April 2021, a cross-sectional online questionnaire hosted on REDCap was administered through access links sent to WhatsApp support groups for HIV PrEP users and attendees of two primary healthcare clinics and two mobile facilities in the Eastern Cape and Gauteng provinces aged between 18-49 years. Participants either self-completed the questionnaire or received support from a research assistant. We used a CLOGIT model for the initial analysis and latent class model (LCM) to establish class memberships with results displayed as odds ratios and probabilities.

We enrolled 496 individuals, the majority were female (69%) and <30 years (74%). About 29% reported previous STI treatment and 20% reported current STI symptoms.

The LCM showed two distinct groups within the respondent sample with different preferences for STI care. The first group comprising 68% of participants showed a strong preference for self-sampling compared to sampling by a healthcare professional (HCP) [OR 2.32; 95%-CI (1.79-3.00)] and viewed no sampling as similar to HCP sampling [OR 1.08; 95%-CI (0.92-1.26)]. There was a lower preference to receive results within 4 hours versus 2 hours [OR 0.63; 95%-CI (0.51-0.77)] and the later was viewed as equal to the receipt of results in 1-7 days by SMS or online [OR 1.03; 95%-CI (0.88–1.21). A clinic follow-up appointment for treatment was less preferable than same-day treatment [OR 0.78; 95%-CI (0.63–0.95)] while treatment from a local pharmacy was viewed with equal preference as same-day treatment [OR 1.16; 95%-CI (1.04-1.29)]. Contact slip from index patient [OR 0.86; 95%-CI (0.76-0.96)] and HCP-initiated partner notification [OR 0.63; 95%-CI (0.55-0.73)] were both less preferable than expedited partner treatment (EPT). The second group included 32% of participants with a much lower preference for self-sampling compared to sampling by HCP [OR 0.55; 95%-CI (0.35–0.86)]. No sampling was not significantly different to HCP-sampling [OR 0.85; 95%-CI (0.64-1.13)]. There was a strong preference for a 4-hour wait than a 2-hour wait for results [OR 1.45; 95%-CI (1.05-2.00)]. There was no treatment option that was significantly different from the others, however there was a strong preference for HCP-initiated partner notification than EPT [OR 1.53; 95%-CI (1.10-2.12)]. Participants were more likely to be members of group 1 than group 2 if they were aged 25-49 years compared to 18-24 years (p=0.001) and receive care from a rural compared to urban facility (p=0.011). Employed individuals were more likely to be in group 2 than group 1 (p=0.038).

**Conclusions:** Our results suggest that health service users preferred to undergo STI testing prior to treatment but there were subgroups who differed on how this should be done. This highlights the need for STI care to be flexible to accommodate different patient choices.

## Introduction

Sexually transmitted infections (STI) present a major global public health problem, especially in women. The common, curable STIs chlamydia, gonorrhoea, syphilis and trichomoniasis are all associated with a higher risk of HIV acquisition and transmission [1]. Complications of STIs resulting in serious morbidity also include poor sexual and reproductive health (SRH) outcomes such as infertility, ectopic pregnancy, poor pregnancy outcomes, congenital infections and cancer [1], most of which are preventable with early identification and treatment.

Whilst STIs affect individuals of all ages, adolescents and young people are disproportionately affected. High rates of STIs have been identified amongst young people in sub-Saharan Africa, with South African household studies reporting prevalence rates of any curable STI of 14% among 15 – 24 year olds [2], and prevalence rates of gonorrhoea of 3.2% and 6.5% and chlamydia of 11.7% and 15.6% [2, 3]. STI prevalence and incidence is also particularly high among young women receiving HIV Pre-Exposure Prophylaxis (PrEP) [4, 5], with studies reporting chlamydia prevalence ranging from 8.0% to 20.6% and gonorrhoea from 1.4% to 8.9% among women aged 15 – 24 years in South Africa. [4]

Although the scale up of PrEP and the provision of an integrated approach to HIV prevention and SRH provides an opportunity to improve STI control [1], STI diagnosis and management services in sub-Saharan Africa have remained relatively unchanged.

The current standard of care in South Africa remains syndromic management in which individuals presenting with STI symptoms receive treatment based on national algorithms [6].The drawback of this approach is that not all symptoms suggestive of STIs are actually due to STIs. For example, in a South African STI surveillance study, the commonest causes of vaginal discharge syndrome were bacterial vaginosis and candidiasis, and 19% of clinic presentations with vaginal discharge did not have a detectable STI pathogen [7, 8]. Furthermore, the majority of people with STIs are asymptomatic, hence will remain undiagnosed and untreated using the syndromic approach [2, 9, 10].

The World Health Organization (WHO) has identified better diagnosis, treatment and partner services for populations at high and ongoing risk of acquiring STIs as key strategies to achieving global targets to ending STI epidemics as public health concerns by 2030 [1]. The South African National Strategic Plan (NSP) for HIV, TB and STIs (2017-2022) highlights the need to improve the detection and management of asymptomatic STIs with increased laboratory support and use of point-of-care (POC) testing [11] and national STI management guidelines make provision for the implementation of asymptomatic STI screening strategies for adolescent girls and young women accessing SRH services [6]. However, implementation of these approaches is limited.

Even with a diagnostic approach, challenges with return for results and treatment provision exist. An estimated 36 – 40% of positive cases of STIs did not receive their results or treatment in one study implementing point of care STI testing in Zimbabwe [12]. Challenges with partner notification and treatment among young people also hinder STI control [13]. In addition, limited knowledge on STIs, long clinic waiting times, accessibility of services, poor integration and services that are not orientated to youth are cited as barriers to obtaining STI and SRH services [14].

Evidence is needed to inform the revitalisation of STI services for young people in sub-Saharan Africa and the transition from syndromic STI care to an aetiology-based approach that takes advantage of the recent advances STI diagnostics and effectively integrates these within existing health services. Furthermore, user perspectives and input into programme design are critical to ensure services are youth friendly and meet the needs and preferences of young people.

We aimed to ascertain health service users’ preferences for diagnosis and treatment of STIs, receipt of results, treatment and partner notification among participants using discrete choice experiments (DCEs).

## Methods

### Study setting

Wits Reproductive Health and HIV Institute implements a PrEP implementation science project (Project PrEP) in four clusters in South Africa. The goal of this project is to inform the introduction of oral PrEP for adolescent girls and young women in South Africa. Two of the clusters were identified as sites for this study. The study included individuals who receive care from the fixed and mobile units of urban and peri-urban primary health care clinics in Tshwane sub-district in Gauteng Province and Nelson Mandela A in Eastern Cape Province of South Africa respectively using a combination of unassisted and research assistant-supported online surveys. In recent studies, the prevalence of any STI amongst pregnant women in the Tshwane sub-district was about 40% [15] and 22.9% amongst women attending a rural community clinic in the Eastern Cape [16]. The antenatal HIV prevalence in Tshwane districts and Nelson Mandela Bay were estimated at 28.7% and 29.7% % respectively [17].

### Study design

The study employed an analytic cross sectional design with a DCE. A DCE is a quantitative method used to measure the strength of these preferences and trade-offs and is grounded in random utility theory (RUT), an economics-based theory which postulates that individuals make rational decisions and that goods are described by their characteristics [18]. Further RUT postulates that individuals are likely to select a service based on maximum utility. Using DCE allows for potential service users to state their preferences given hypothetical scenarios, goods or services that are presented to them even when the options are not yet available [19]. The responses given are then used to infer value placed on each attribute.

Participants were required to complete a questionnaire in which they were asked to imagine that they were being offered a test for STI during a clinic attendance prior to answering a series of questions (S1 Table) in which they were required to choose service preferences offered by either Clinic A or Clinic B. The options were described according to a number of characteristics (known as ‘attributes’) such as how and when treatment for the STI is given. Each attribute also had a number of ‘levels’, such as ‘at the clinic the same-day’, ‘at the clinic during a follow up appointment’ or ‘from a place near you such as a local pharmacy’.

The study is reported according to the Strengthening the Reporting of Observational Studies in Epidemiology (STROBE) guidelines.

### DCE instrument design

#### Choice of attributes and levels

The DCE attributes relating to STI sample collection, diagnosis, receipt of results, receiving treatment and notification of partners were informed by a review of relevant literature [10, 20-23]. This informed the study matrix of attributes and levels as described in Table 1 for each service delivery consideration (attributes). Attributes were considered as characteristics of a service delivery model for STI care and treatment. There were four attribute and each had 3-4 ‘levels’. Levels are the value of scenarios that each attribute can take on. The levels assigned represent both standard of care and alternatives listed in literature and these were refined through multiple discussions with, individuals who may represent the demographic of the participants, study team/collaborators, methodologists and experts in the field.

**Table 1.** Discrete Choice Experiments attributes and levels

| Attributes | Levels | Description |
| --- | --- | --- |
| **How the STI testing sample is taken** | No sample is taken, care is based only on presenting symptoms | No samples are taken – you are given treatment just based on whether you have symptoms or not |
|  | You self-sample at the clinic | There are different types of samples – urine for men and vaginal swab for women [Display swab]. After being provided with instructions, you could take the sample yourself at a private place at the clinic |
|  | By a health care professional at the clinic | The provider takes a sample when he/she is examining you |
| **How and when you are told if you have an STI** | The same day after a 2 hour wait at the clinic by a health care professional | This may be the shortest time you will spend in the clinic in total because diagnosis is based on symptoms only, no samples are taken so the provider will not know whether you really have an infection or not and will also not know which infection/s you have |
|  | The same day after a 4 hour wait at the clinic by a health care professional | You will spend a longer time in the clinic because samples will be taken and tested at clinic so you will definitely know if you have an infection and what it is |
|  | In 1-7 days' time via a secure online website | You may spend about 2 hours in the clinic because samples will be taken but it will not be tested at the clinic but rather, they will be sent away for testing so your results will not be available on the same day. You will be given a secure code at the clinic and a website address to check your results privately. If you have an infection, you may still have to go back to the clinic for treatment. |
|  | In 1-7 days' time via text message (SMS) | You may spend about 2 hours in the clinic because samples will be taken but it will not be tested at the clinic but rather they will be sent away for testing so your results will not be available on the same day. You will receive a text message when your results are ready which will either say [Your Results Are Ready – No Follow-up Visit Required] or [Your Results Are Ready – Please present at your preferred clinic as soon as possible]. If you have an infection you will have to go back to the clinic for treatment. |
| **How and when treatment for your STI is given if required** | From a place near you such as a local pharmacy | Medication can be picked up from a local pharmacy |
|  | At the clinic during a follow-up appointment | If you receive a SMS requiring you to revisit the clinic or if the online system indicates you have an infection, you will be required to come back to the clinic to collect your treatment |
|  | At the clinic the same day | If you are being treated according to your symptoms or not or if are being tested and the test in being done at the clinic on the same day |
| **How your partner(s) is / are notified if you have an STI** | You notify your partner using a notification slip | We will give you a slip to take to your partner/s. The slip will inform your partner to attend the clinic for diagnosis and/or treatment. |
|  | Your partner(s) is / are notified directly by a health care professional | If you prefer, the provider can contact your partner and invite them to the clinic for testing and treatment, without mentioning you |
|  | Expedited partner treatment | If you are being treated for an infection you will be given treatment to give to your partner/s for the same infection. |

The four attributes and their respective levels comprising are described in Table 1

There was a total of 108 possible combinations of the STI attributes and levels that were selected for the study (3 × 3 × 3 × 4), resulting in 5778 total combinations [(108 x107)/2] across two scenarios A or B. This number of combinations are too many to present to the participants, hence the DCE instrument was designed using a D-efficient approach with 12 choice tasks (questions) using Ngene software [24]), ensuring that preferences for each of the attribute levels could be independently assessed.

Participants were also asked to provide information on sociodemographic and risk factors including age (18-24/25-29/≥30), gender (male/female), access to mobile phone (Yes/No), access to internet (Yes/No), employment status (part time or full time/unemployed/other), distance from preferred clinic in Km, relationship status (married/in a stable relationship/casual partners/single, no partners, casual relationships only, other), number of current sex partners (0/1/2-4/>4), sex with a partner whose HIV status is unknown (Yes/No), unprotected sex in last year without a condom (Yes/No), Sex with partners who are HIV positive (Yes/No/Unsure), sex under the influence of alcohol or drugs (Yes/No), Previous STI treatment (Yes/No), current STI symptoms (Change in Vaginal Discharge [smell, colour, quantity]/lower abdominal pain/Pain during sex/genital sores/pain when passing urine/urethral discharge [“drops”] (Yes for any in the list, otherwise No). The complete questionnaire is available in the S1 Table.

### Procedures and data collection

The DCE questionnaire was administered between 1 March 2021 and 20 April 2021. Piloting of the study recruitment and data collection processes was undertaken prior to study implementation and informed the final recruitment and data collection strategies. Participants aged 18-49 years old were recruited from study sites using the following approaches:

#### i) Direct link to questionnaire received by participants

Potential participants accessed or received a link to the online questionnaire through either clinic WhatsApp groups (support groups for PrEP users moderated by peer navigators or demand creation officers), via an existing participant or study site client who shared the study link they had received, through a poster advertisement at the study site, or via a research assistant who sent study links to potential participants identified through the clinic database and contacted telephonically prior to sharing the questionnaire link to assess interest in the study. A recorded voice message guided participants through the informed consent process and the questionnaire.

#### ii) Recruitment at the healthcare facility

Individuals attending either the mobile or fixed clinic facilities at the study sites were recruited to the study by a research assistant, who then supported the participant to complete the study questionnaire.

Regardless of the approach used for participant recruitment, there were two methods of data collection; i) online questionnaire completion without research assistant support henceforth referred to as Data Collection (DC)-1 ii) Online questionnaire completion with research assistant support henceforth referred to as DC-2.

All participants received the equivalent of USD14 in the local currency taking into account internet data costs and time spent on the study.

Ethical approval from the study was obtained from the Human Research Ethics Committee of the University of the Witwatersrand, South Africa (ref: M200445) and the Research Governance and Ethics Committee of the Brighton and Sussex Medical School, UK (ref: ER/BSMS9B5G/3).

Determining sample sizes in advance of conducting DCEs is difficult, because the questionnaire design is usually not known at the study’s outset. However, as it has been suggested that a reasonable sample size is 300, we aimed to recruit at least 500 participants with complete answers to allow us examine predictors of group membership in a planned latent class modelling [25].

### Statistical analysis

Descriptive characteristics of the study participants were tabulated according to the data collection method used.

The results are presented as odds ratios (ORs) relative to the relevant base category and show the probabilities of uptake of any of the other categories. Reported standard errors are adjusted in all instances to account for the potential clustering in participant responses.

We used the conditional logit (CLOGIT) model to analyse the DCE [26]. Unlike in standard logistic regression, the results from CLOGIT models are ‘conditional’ on the information relating to all the choice options as this information is grouped before analysis. In this sense, CLOGIT models are akin to matched case–control designs as they investigate the relationship between a choice (case), options that were not chosen (controls), and a set of predictive factors (attribute levels).

Although the CLOGIT model is recommended to be used for the initial analysis [27], it produces results for the ‘average’ individual, meaning that no allowance is made for the possibility that different groups of people within the sample (e.g., different age groups) might have varying preferences—this is known as ‘preference heterogeneity’.

As an alternative to the CLOGIT approach, we also analysed the results using a latent class model (LCM), as it simultaneously relaxes the independence of irrelevant alternatives assumption and allows potential preference heterogeneity to be examined. LCMs are recommended if groups of respondents with similar preferences are anticipated.

LCMs assume there are subgroups of individuals (classes) with similar preferences, and that the likelihood of class membership can be related to observed variables. The potential predictors of class membership in this analysis were gender, age (as a categorical variable), current STI symptoms (yes; no), previous STI treatment (yes; no), facility location (urban; rural, other), employment (not employed, part or fulltime employment; other), method of data collection (no research assistant support, research assistant support provided)

The number of classes in the LCM was based on minimisation of Akaike’s information criterion (AIC) and the production of stable/meaningful standard errors. The CLOGIT and LCM analyses were undertaken using Stata version 16 [(StataCorp. Release 16.1. College Station (TX)] and NLOGIT 5 [Econometric Software. NLOGIT 5. Plainview (NY)] respectively.

## Results

### Characteristics of participants

Of the 443 participants who clicked on the online questionnaire link for DC-1, (no assistance provided for questionnaire completion), 291 completed the full questionnaire. The remaining individuals discontinued prior to providing informed consent, hence were not included in the analysis. Of the 213 individuals who received support to complete the online questionnaire (DC-2), 205 provided consent and completed the full survey. In total, 496 individuals completed the questionnaire across the two data collection methods. About 367 (74%) of the participants were <30 years old with a median of 25 years (IQR 22, 29) and there was no age difference between the two groups by method of data collection. The majority of the participants were female (69%) with no gender difference by method of data collection. There was evidence of high-risk sexual behaviour amongst recruited participants with 144 (29%) having previously been treated for STIs and 101 (20%) reporting current STI symptoms. Those in DC-2 were more likely to have reported their employment status as ‘other’ (p<0.001), less likely to have reported condomless sex in the last year (p<0.001) and more likely to be users of the clinic facilities selected for recruitment of participants (p<0.001) than those in DC-1. (Table 2)

**Table 2:**
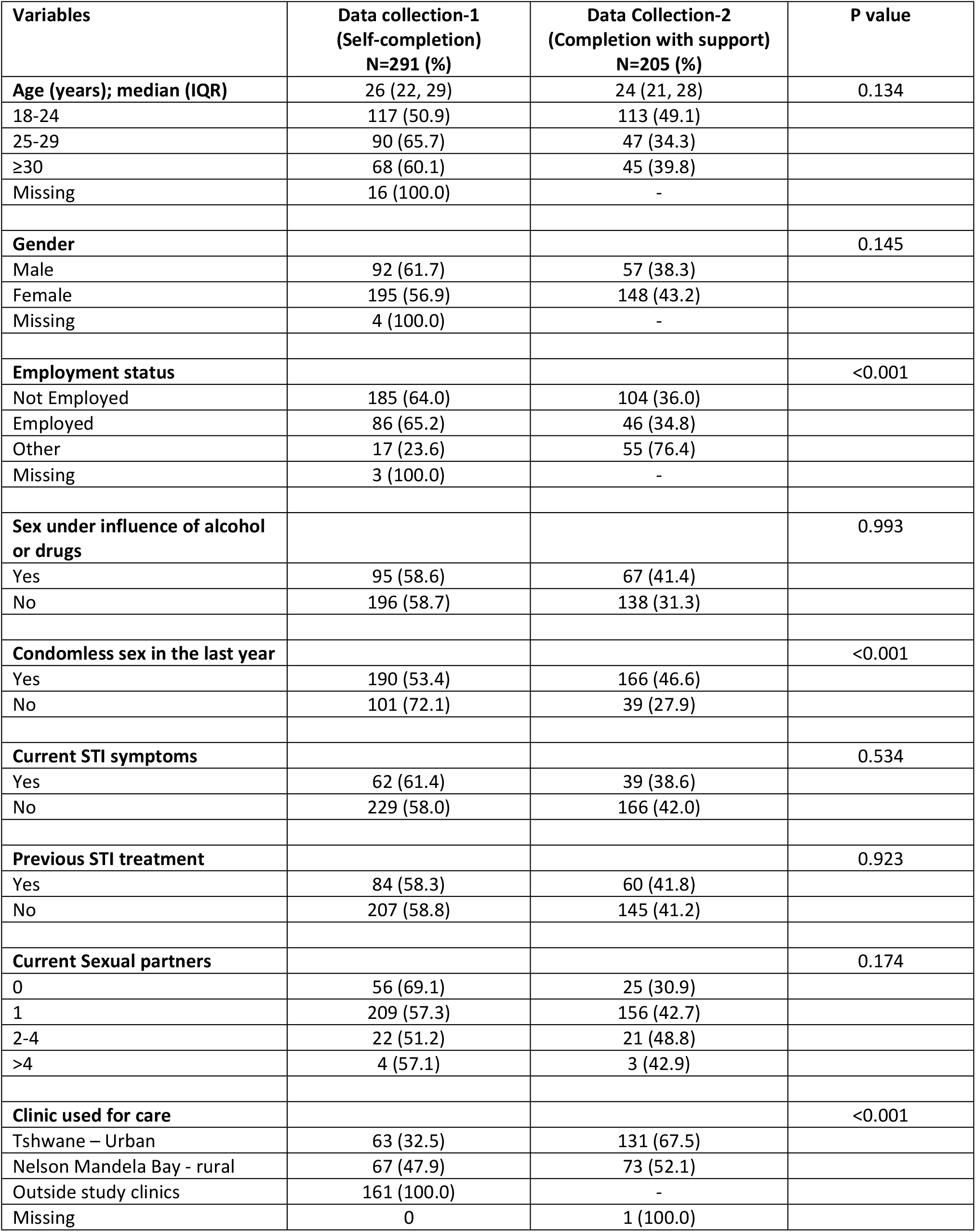
socio-demographic and sexual risk behaviour by data collection methods.

| **Variables** | **Data collection-1**<br>**(Self-completion)**<br>**N=291 (%)** | **Data Collection-2**<br>**(Completion with support)**<br>**N=205 (%)** | **P value** |
| --- | --- | --- | --- |
| **Age (years); median (IQR)** | 26 (22, 29) | 24 (21, 28) | 0.134 |
| 18-24 | 117 (50.9) | 113 (49.1) |  |
| 25-29 | 90 (65.7) | 47 (34.3) |  |
| ≥30 | 68 (60.1) | 45 (39.8) |  |
| Missing | 16 (100.0) | - |  |
| **Gender** |  |  | 0.145 |
| Male | 92 (61.7) | 57 (38.3) |  |
| Female | 195 (56.9) | 148 (43.2) |  |
| Missing | 4 (100.0) | - |  |
| **Employment status** |  |  | <0.001 |
| Not Employed | 185 (64.0) | 104 (36.0) |  |
| Employed | 86 (65.2) | 46 (34.8) |  |
| Other | 17 (23.6) | 55 (76.4) |  |
| Missing | 3 (100.0) | - |  |
| **Sex under influence of alcohol or drugs** |  |  | 0.993 |
| Yes | 95 (58.6) | 67 (41.4) |  |
| No | 196 (58.7) | 138 (31.3) |  |
| **Condomless sex in the last year** |  |  | <0.001 |
| Yes | 190 (53.4) | 166 (46.6) |  |
| No | 101 (72.1) | 39 (27.9) |  |
| **Current STI symptoms** |  |  | 0.534 |
| Yes | 62 (61.4) | 39 (38.6) |  |
| No | 229 (58.0) | 166 (42.0) |  |
| **Previous STI treatment** |  |  | 0.923 |
| Yes | 84 (58.3) | 60 (41.8) |  |
| No | 207 (58.8) | 145 (41.2) |  |
| **Current Sexual partners** |  |  | 0.174 |
| 0 | 56 (69.1) | 25 (30.9) |  |
| 1 | 209 (57.3) | 156 (42.7) |  |
| 2-4 | 22 (51.2) | 21 (48.8) |  |
| >4 | 4 (57.1) | 3 (42.9) |  |
| **Clinic used for care** |  |  | <0.001 |
| Tshwane – Urban | 63 (32.5) | 131 (67.5) |  |
| Nelson Mandela Bay - rural | 67 (47.9) | 73 (52.1) |  |
| Outside study clinics | 161 (100.0) | - |  |
| Missing | 0 | 1 (100.0) |  |

### STI care preferences

#### Preference of the ‘average’ individual

The CLOGIT model produced results showing the STI preferences of the ‘average’ individual (Table 3). This showed a strong preference for self-sampling [OR 1.49 (95% CI: 1.32-1.68)] with no significant difference between no sampling, in which treatment is based on syndromes, and healthcare professional (HCP) sampling. There was a much lower preference for having to wait 4 hours in the clinic to get results on the same day compared to a 2 hour wait [(OR: 0.82 (95% CI: 0.73-0.92)]. However, there was no difference in preference between waiting for 2 hours to get results on the same day and getting the results in 1-7 days by either SMS or secure online portal. There was a slightly lower preference for a clinic follow up for treatment of STI compared to same day treatment [OR 0.91 (95% CI 0.81-1.01; p=0.077)]. There was no significant difference between a preference for same day clinic treatment and treatment at a local pharmacy. There was a lower preference for partner notification by the index patient [(OR 0.93 (95%-CI 0.87-0.99)] or provider-initiated [(OR 0.83 (95%-CI 0.77-0.89)] compared to expedited partner treatment (EPT).

**Table 3:** STI testing preferences of the ‘average’ individual.

| Variables | Odds ratio (95% CI) | P value |
| --- | --- | --- |
| **How STI samples should be taken** |  |  |
| HCP | 1 |  |
| Self | 1.49 (1.32-1.68) | <0.001 |
| No sampling | 1.01 (0.92-1.11) | 0.845 |
| **How and when results are given** |  |  |
| Same day after 2hrs | 1 |  |
| Same day after 4hrs | 0.82 (0.73-0.92) | 0.000 |
| 1-7 days' time via SMS/online | 1.05 (0.95-1.16) | 0.301 |
| **How and when treatment is given** |  |  |
| Same day in clinic | 1 |  |
| Clinic follow-up on appointment | 0.91 (0.81-1.01) | 0.077 |
| Pharmacy near patient | 1.06 (0.99-1.13) | 0.110 |
| **How partners are notified of an STI** |  |  |
| Expedited partner treatment | 1 |  |
| Contact slip from index patient | 0.93 (0.87-0.99) | 0.028 |
| HCP initiated notification | 0.83 (0.77-0.89) | 0.000 |

#### Latent class model

The latent class model identified two groups, with 68% and 32% of participants likely to be in groups 1 and 2 respectively (Table 4).

**Table 4:** STI testing preferences-Results from the latent class model.

| Variables | Group 1 (68%)<br>Self-led |  | Group 2 (32%)<br>HCP-led |  |
| --- | --- | --- | --- | --- |
|  | Odds ratio (95% CI) | P value | Odds ratio (95% CI) | P value |
| **How STI samples should be taken** |  |  |  |  |
| HCP | 1 |  | 1 |  |
| Self | 2.32 (1.79-3.00) | <0.001 | 0.55 (0.35 – 0.86) | 0.008 |
| No sampling | 1.08 (0.92-1.26) | 0.368 | 0.85 (0.64-1.13) | 0.265 |
| **How and when results are given** |  |  |  |  |
| Same day after 2hrs | 1 |  | 1 |  |
| Same day after 4hrs | 0.63 (0.51- 0.77) | <0.001 | 1.45 (1.05- 2.00) | 0.023 |
| 1-7 days' time via SMS/online | 1.03 (0.88 – 1.21) | 0.689 | 1.07 (0.81 – 1.41) | 0.640 |
| **How and when treatment is given** |  |  |  |  |
| Same day in clinic | 1 |  | 1 |  |
| Clinic follow-up on appointment | 0.78 (0.63 – 0.95) | 0.012 | 1.27 (0.92 – 1.76) | 0.149 |
| Pharmacy near patient | 1.16 (1.04-1.29) | 0.006 | 0.86 (0.72 – 1.04) | 0.121 |
| **How partners are notified of an STI** |  |  |  |  |
| Expedited partner treatment | 1 |  | 1 |  |
| Contact slip from index patient | 0.86 (0.76-0.96) | 0.010 | 1.10 (0.92-1.32) | 0.285 |
| HCP initiated notification | 0.63 (0.55-0.73) | <0.001 | 1.53 (1.10-2.12) | 0.011 |

Those in group 1 had a strong preference for self-sampling compared to HCP sampling [(OR 2.32; (1.79-3.00) p<0.001] with no significant difference between HCP sampling and no sampling (syndromic management). There was a much lower preference for a 4-hour wait for results compared to a 2-hour wait [(OR 0.63; (95% CI 0.51-0.77) p<0.001]. However, there was no significant difference in preference between a 2-hour wait for same day results and getting the results in 1-7 days by either SMS or secure online portal. There was a lower preference for a clinic follow up appointment for treatment than same day treatment [OR 0.78; (95% CI 0.63 - 0.95) p=0.012]. However, there was a strong preference for receiving treatment from a local pharmacy compared to waiting 2 hours for same day treatment [OR 1.16; (95% CI 1.04-1.29) p=0.006]. There was a much lower preference for partner notification by the index patient [(OR 0.86; (0.76-0.96) p=0.010] or HCP-initiated [(OR 0.63; (0.55-0.73) p<0.001)] compared to EPT. We referred to group 1 members as those preferring a self-led service

Participants in group 2 had a much lower preference for self-sampling compared to HCP sampling [OR 0.55; (95% CI 0.35-0.86) p=0.008]. There was no statistically significant difference between no sampling and HCP sampling. There was a strong preference for waiting 4 hours for same day results compared to 2 hours [OR 1.45; (1.05-2.00) p=0.023]. There was no significant difference between receiving results in 1-7 days by SMS/online compared to 2 hours on the same day with both being less preferred options. There was some evidence that a clinic follow up for treatment was preferred to same day treatment although this did not reach statistical significance [OR 1.27 (95% CI 0.92 – 1.76) p=0.149]. Receiving treatment from a local pharmacy was less preferred compared to same day treatment but this did not reach statistical significance [OR 0.86 (95% CI 0.72 −1.04) p=0.121]. There was a strong preference for partner notification by an HCP compared to EPT [OR 1.53 (95% CI 1.10-2.12) p=0.011] and no significant difference between partner notification by index patient and EPT. We refer to group 2 participants as those preferring an HCP-led service.

Participants were more likely to prefer a self-led service to an HCP-led service if they were aged 25-49 years compared to 18-24 years (p=0.001) and to receive care from a rural rather than an urban facility (p=0.011). Employed individuals were more likely to prefer an HCP-led service to a self-led service (p=0.038). Gender, current STI symptoms, previous treatment of STIs and method of data collection were not predictive of class memberships.

## Discussion

We conducted a DCE to investigate young people’s preferences for the management of STIs in primary healthcare. Our results showed that the ‘average’ individual prefers to self-sample, receive their result within 2 hours, may not want to come back to the clinic on a different day for treatment and would prefer EPT to other forms of partner notification. However, the latent class model showed that there were two groups of individuals with different STI service delivery preferences. About two-thirds of individuals, whom we have classified as preferring a self-led service, would prefer to take their own genital sample, be informed of their results within 2 hours on the same day or 1-7 days later by SMS/online, receive treatment from a pharmacy that is local to them and use EPT. These individuals were more likely to be older than 25 years and receive care from rural healthcare facilities.

The remaining one-third of individuals, whom we have classified as preferring HCP-led service showed a preference for HCP sampling and would be willing to wait up to 4 hours in a clinic to get their results, with some indication that they may not mind returning to the clinic for treatment and would prefer the HCP to undertake partner notification if required. These individuals were more likely to be younger than 25 years, employed and receive their care from urban healthcare facilities.

The South African PrEP guidelines recommend either syndromic or aetiology-based STI care in individuals at high risk of HIV acquisition who require PrEP. However, aetiology-based STI care is not routinely available in the public health programme except as part of implementation research in certain facilities. The service currently available for STI care in health facilities differs from the preferences of service users based on our study results. Our results show that individuals need to be presented with choices as a one size fits all approach will not meet the needs of all individuals.

Individuals indicated a preference for providing genital samples for the diagnosis of STIs either through a self-taken swab or by a HCP taking the sample, with the majority preferring the former approach. This demonstrates that service users want to be tested for the presence of an actual STI prior to being treated. Other studies in South Africa have also demonstrated that the self-taken swab is acceptable and feasible [2, 28]. In one of the studies, nearly two-thirds of individuals preferred self-taken swabs, and the remaining one-third either preferred HCP-taken swabs or expressed no preference [28]. We observed that younger individuals who receive their care from rural clinics showed a preference for getting their results within 2 hours on the same day or by SMS in 1-7 days, with the later choice being available to those who wanted a diagnostic test. There was a preference for same day treatment or treatment from a local pharmacy with a clinic follow-up being less preferred. This would suggest that an aetiology-based approach is preferred by this group if results can be provided within 2 hours on the same day or substituted with the convenience of receiving treatment from a local pharmacy. The unwillingness of patients to wait >2hours for results and treatment is consistent with another study that evaluated the feasibility of same day test and treatment using CT/NG GeneXpert in a clinic in a high-income setting. The authors concluded that the processing time for GeneXpert was too long to be used as a point of care test [29]. In a South African study, it was estimated that patients had to wait for a mean of nearly 5 hours for STI POC diagnostic testing using GeneXpert CT/NG assay which is unlikely to be acceptable to healthcare users [20]. The challenges of using GeneXpert CT/NG as an STI POC diagnostic tool was further highlighted by a recent study in Zimbabwe in which Youth were unwilling to wait 90 minutes for their results [30].

About one-third of those who participated in the DCE indicated they would prefer to wait for up to 4 hours if it meant that they received diagnostic testing for their STIs, with weak evidence of a preference to return to the clinic for treatment during a follow up appointment. The likelihood of receiving same day treatment would be high in this group as they were willing to wait 4 hours for aetiology-based diagnosis, which might be the reason for a lack of a statistical difference between the three treatment options presented. The two groups of individuals in this study differed in how they would want their sexual partners to be notified if they were diagnosed with an STI. Two-thirds of individuals had a strong preference for EPT, whilst a third preferred an HCP-initiated notification. EPT has been shown to be acceptable to young South African women and their partners [10, 31] and resulted in a decrease in STIs in a follow up test amongst those who accepted EPT [10]. Some of the reasons provided for this preference included that partners may agree to visit a clinic but would not do so, hence they will prefer to witness their partners taking the medication, partners may be too busy to visit the clinic or may refuse [31]. Some patients may not be comfortable discussing STIs with sexual partners, hence the option of HCP-initiated partner notification should continue to be offered in the healthcare setting.

This DCE indicates that users of an STI service will prefer an aetiology-based STI management approach and models of STI care that allow them to obtain their results and treatment in a prompt and convenient manner. This would mean making available a number of treatment options for aetiology-based STI care that includes same day clinic treatment as well as community treatment. The availability of POC STI technologies that provide results for CT, NG and trichomonas vaginalis in 30 minutes [22, 23] puts this model of aetiology-based STI care within reach, potentially allowing an episode of care to be completed within 2hours of the patient arriving in the clinic. Aetiology-based STI care has the advantage of detecting STIs in asymptomatic individuals who would otherwise remain undiagnosed and untreated. It could also prevent overtreatment of people with STI symptoms with antibiotics for STIs which they do not have, hence contribute to reducing the burden of antimicrobial resistance through both good antibiotics stewardship and reduction in STI burden [32].

Effective STI care is a good HIV prevention strategy as STIs are associated with increased HIV transmission and acquisition [33]. Hence aetiology-based STI care should be integrated within PrEP programmes targeting individuals at high risk of HIV acquisition, such as adolescent girls and young women, sex workers and men who have sex with men. STIs also result in substantial burden of disease such as pelvic inflammatory disease, ectopic pregnancy, infertility, still births, cancers as well as significant social harm [34]. This supports the argument to strengthen the health system to provide aetiology-based STI care to anyone requesting STI care regardless of their risk group.

Our study has a few limitations. Firstly, participants were presented with a hypothetical scenario in which they were asked to imagine they were having an STI test. It is possible that some participants could struggle to determine their preference for STI care (e.g. self-testing, EPT) which they have never experienced, however, there is growing evidence that DCEs predict actual behaviour [35, 36]. Secondly, we used two data collection approaches in which participants either completed the questionnaire without assistance or received support from a research assistant to complete the questionnaire. The individuals in the two approaches differed by their employment status and the facilities they attended for care, with employment status being predictive of class membership in the latent class model, hence this could have biased our results. As only individuals who had access to mobile phones participated in the survey, our results may not be generalizable to those individuals who may not easily access a mobile phone. It is reassuring that internet access via a mobile device is growing every year in South Africa and is estimated to be 60.7% in 2021 [37]. As a result, it is likely that mobile technologies will play an increasing role in facilitating access to healthcare.

Our study also has a few strengths. Recruitment through WhatsApp, social media, rural and urban clinic settings meant we were able to reach different categories of individuals including those with risk factors for STIs as shown in the baseline characteristics of the participants.

In summary, our results suggest that all individuals preferred to undergo STI testing prior to treatment but there were subgroups who differed on how this should be done. The majority of individuals preferred a self-led service, that requires them to spend as little time as possible in the clinic either for results or treatment and would prefer for some aspects of their care to happen outside of clinic facilities. They also showed a strong preference for EPT. The other group, although smaller, preferred their care to be HCP-led and clinic-based and would be willing to wait longer for results and return for treatment during a follow up appointment. They would also prefer their partners to be notified by the HCP. This highlights the need for a range of STI care options to be available to young people, who comprise the majority of our participants, as one size does not fit all.

## Data Availability

We will make the data available either as part of the final submission if the article is accepted or it will be hosted on the Institutional website. We will communicate which of these options will apply on acceptance of the article.
Supplementary materials submitted as part of the main manuscript to PLOS MED

## Acknowledgement

We thank the South African National Department of Health for their support. We also thank our patients for their sacrifices and willingness to support research in order to improve lives

## Funding

This research was commissioned by the National Institute for Health Research (NIHR) Global Health Policy and Systems Research programme using UK aid from the UK Government. The views expressed in this publication are those of the author(s) and not necessarily those of the NIHR or the Department of Health and Social Care (Grant No. NIHR130250).

PS is supported by Consortium for Advanced Research Training in Africa (CARTA). CARTA is jointly led by the African Population and Health Center and the University of the Witwatersrand and funded by the Carnegie Corporation of the New York (Grant No. B 8606.R02, SIDA (Grant No: 54100029), the DELTAS Africa Initiative (Grant No. 107768/Z/15/Z). The DELTAS Africa Initiative is an independent funding scheme of the African Academy of Sciences (AAS)’s Alliance for Accelerating Excellence in Science in Africa (AESA) and supported by the New Partnership for Africa’s Development Planning and Coordinating Agency (NEPAD Agency) with funding from the Wellcome Trust (UK) and the UK government. The statements made and views are solely the responsibility of the Fellow.

## Transparency declarations

CI has received conference support and research grants from Gilead Sciences. All other authors have nothing to declare.

## Notes

### Competing Interest Statement

I have read the journal's policy and the authors of this manuscript have the following competing interests: CI has received conference support and research grants from Gilead Sciences. All other authors have declared that no competing interests exist

